# UK healthcare professionals’ attitudes towards the introduction of varicella vaccine into the routine childhood vaccination schedule and their preferences for delivery

**DOI:** 10.1101/2023.09.02.23294950

**Authors:** Susan M. Sherman, Charlotte Allerton-Price, Nicola Lingley-Heath, Jasmine Lai, Helen Bedford

## Abstract

**Background:** Varicella (chickenpox) is a highly contagious disease caused by the varicella-zoster virus. Although typically a mild disease, varicella can cause complications leading to severe illness and even death. Safe and effective varicella vaccines are available. The Joint Committee on Vaccination and Immunisation is planning to review the evidence regarding the introduction of varicella vaccine into the UK’s routine childhood immunisation schedule.

**Objectives:** To explore UK healthcare professionals’ (HCPs) knowledge and attitudes towards varicella vaccination, its introduction to the UK routine childhood immunisation schedule, and their preferences for how it should be delivered.

**Design:** We conducted an online cross-sectional survey exploring HCPs’ attitudes towards varicella, varicella vaccine, and their preferences for delivery of the vaccine between August and September 2022.

**Participants:** 91 HCPs working in the UK (96.7% female, 3.3% male, mean age 48.7 years).

**Results:** General vaccine attitudes in this group were very positive. Gaps in knowledge about varicella were revealed: 21.0% of respondents disagreed or were unsure that chickenpox can cause serious complications, while 41.8% were unsure or did not believe chickenpox was serious enough to vaccinate against. After receiving some basic information about chickenpox and the vaccine, almost half of the HCPs (47.3%) in our survey would prefer to administer the varicella vaccine combined with MMR.

**Conclusions:** Given the positive influence of HCPs on parents’ decisions to vaccinate their children, it is important to understand HCPs’ views regarding the introduction of varicella vaccine into the routine schedule. Our findings highlighted areas for training and HCPs’ preferences which will have implications for policy and practice should the vaccine be introduced.

## INTRODUCTION

Varicella (chickenpox) is a highly contagious disease caused by the varicella-zoster virus (VZV) (Gonzalez-Martin, 2019). Common symptoms include an itchy rash with small fluid filled blisters (vesicles), fever, fatigue, loss of appetite and malaise which can last up to two weeks. Viral transmission occurs through direct contact with an infected person, such as respiratory droplets or the fluid from the vesicles. After initial infection, the varicella virus remains latent in the individual’s sensory nerve ganglia where, in 10-20% of cases, it reactivates usually in later life, causing herpes zoster (shingles) (WHO, 2016). Whilst varicella can be contracted at any age, in temperate areas 90% of cases are seen in children before the age of 10 years (Helmuth, et al 2015). In most cases the disease is mild, although serious complications can occur, more commonly in adults particularly pregnant women and in immunocompromised individuals. Other complications occurring at an estimated rate of 2-6% (Rodrigues et al., 2023) include secondary bacterial infections of the skin or soft tissues. Recent chickenpox infection is an important risk factor for invasive Group A streptococcus (iGAS) in children (Zangarini et al., 2023).

Safe and effective varicella vaccines have been available since the 1970s and in routine use in USA since 1995 and Australia since 2005, proving highly successful in reducing disease (Seward et al., 2002; Liese et al., 2008). The vaccine can be administered as a monovalent or a quadrivalent vaccine in combination with the measles, mumps and rubella vaccines (MMRV). In a recent survey of UK parents’ acceptance of varicella vaccine, parents reported a preference for a combination MMRV vaccine, or an additional vaccination visit for a single varicella vaccine over an additional injection at the same vaccination visit as existing vaccines (Sherman et al., 2023). This preference has been borne out in practice in Canada, where the introduction of a combination MMRV vaccine led to higher uptake of varicella vaccine compared with when it was offered separately (MacDonald et al., 2020).

A single-dose of varicella vaccine has been estimated to provide about 80% protection against all infection and almost 100% protection against moderate or severe disease (Marin et al., 2016). In cases where vaccinated individuals do contract varicella, their symptoms are typically milder with fewer complications (Annunziato & Gershon, 2000). Despite this, many countries including the UK currently only recommend varicella vaccination for high-risk groups and individuals in close contact with an immunocompromised individual. Implementation of a routine childhood varicella vaccination programme in UK was considered but not recommended by the Joint Committee on Vaccination and Immunisation (JCVI) in 2010 and this position is currently being reviewed again.

Through epidemiological modelling, the JCVI (2010) predicted that with a high uptake, incorporating varicella vaccine into the routine schedule would see a large reduction in cases of varicella. However, it was also predicted that in the years following its introduction there would be an increase in cases of herpes zoster in adults. This is because it has been hypothesised that vaccinating children would remove or reduce circulating VZV which boosts immunity against herpes zoster; this lack of boosting could lead to an increase in herpes zoster. A shingles vaccine programme for older people been in place in UK since 2013. Furthermore, there has been concern that the reduction in circulating virus would result in an upward shift in the age of acquiring chickenpox infection putting susceptible individuals at risk of disease in adulthood when it is more severe. To avoid these potential consequences of vaccine introduction, it is estimated that a vaccine uptake of above 80% is necessary (WHO, 2014).

A recent study (Sherman et al., 2023) highlighted the trust parents place in advice from the NHS and healthcare professionals (HCPs) when considering vaccination for their children. More than 80% of the 596 parents surveyed, strongly or somewhat agreed that they trusted the advice given on vaccination by practice nurses, health visitors, and their GP. These findings were echoed by the most recent tracking survey of over 1000 parents of children under five years conducted by UK Health Security Agency (UKHSA, 2023). Verger et al., (2022) describes HCPs’ interactions with their patients as “the keystone enabling confidence in vaccination and successful reduction of vaccine hesitancy” (page 2). Through effective communication, HCPs can reassure parents of the safety and benefits of vaccination, addressing any of their concerns so they are able to make informed decisions in their child’s best interests (Verger et al., 2022). Conversely, vaccine-hesitant HCPs may exacerbate or create doubts for parents.

Given the influence HCPs can have on parents’ vaccination decisions, it is important to understand their own views not just as professionals working within the healthcare sector, but also as members of the public and in some cases parents themselves. Building on the work of Sherman et al., (2023), and in anticipation of the JCVI review, this study aims to explore HCPs’ views about the potential introduction of a varicella vaccine into the routine childhood schedule, and what they anticipate parents’ responses to be. This study will also seek HCPs’ insight into any potential barriers or facilitators they consider may have an impact on uptake, as well as which option for delivering the varicella vaccine they would prefer.

## METHOD

### Design

We conducted a cross-sectional survey between 10^th^ August 2022 – 29^th^ September 2022. Participants completed the survey online on the web-based survey tool Qualtrics.

### Participants

Participants (n=91) were recruited through snowball sampling via contacts within the research team, the Royal College of Nursing, and social media pages for example nurses’ Facebook groups. Participants were eligible to take part if they were a healthcare professional, living and working in the UK, who provided advice about or administered childhood vaccines. Of 109 individuals who began the survey, 91 completed it (96.3% completion rate). Although we included 2 attention check questions, no participants failed both and so none were excluded.

### Measures

The questionnaire was developed based on our recent survey of parental attitudes towards the introduction of varicella vaccination (Sherman et al, 2023) and a literature review of vaccine hesitancy in healthcare professionals and varicella vaccination. The final questionnaire is available online (https://osf.io/vejmz/?view_only=2ace6027f9b4499c88059d24a9c3fc63 [peer review link to be updated on acceptance with public link]).

Participants were asked for their demographic details. Questions were asked about: their attitudes towards vaccination generally; knowledge about the chickenpox virus and their perceptions of its risk and seriousness; their own experience of chickenpox; likely parental responses to the introduction of chickenpox vaccine and preferences for how it should be administered; their own preference for how it should be administered and likely barriers and facilitators to parental uptake of the vaccine.

Specifically, we asked participants to report their age, gender, ethnicity, religion, primary healthcare role, the region in which they practice, whether they had children under the age of 18, and if so, whether their children were up to date with their childhood vaccines as per the UK vaccination schedule.

To explore attitudes to vaccination in general, we modified eight items from the WHO SAGE Vaccine Hesitancy Scale (VHS) (Larson et al, 2015; Shapiro et al, 2018) for use in a UK setting and aimed at all participants, not just those with children, to assess attitudes towards childhood vaccines (e.g., reworded from “childhood vaccines are important for my child’s health” to “childhood vaccines are important for a child’s health”). A further item from the VHS was included for parents only (“Generally I do what my doctor or health care provider recommends about vaccines for my child/children”). We also included 9 additional items such as “vaccines are generally safe”, “I believe that healthcare professionals should be up to date with their vaccines to work in the healthcare sector”, “children are vaccinated too early” and “talking with people who are uncertain about vaccination, increases my own uncertainty about vaccination” adapted from Di Martino et al., (2020) and Tomljenovic et al., (2021). Items regarding perceptions of vaccine safety and effectiveness were clarified by including definitions of ‘safe’ (‘means serious side effects are rare’) and ‘effective’ (‘means that most vaccines give good protection’) to ensure consistent interpretation by respondents. Participants were asked to indicate the extent to which they agreed or disagreed with the items on a five-point Likert scale.

We then asked participants to indicate on a five-point Likert scale (from strongly agree to strongly disagree) their agreement with statements: “Chickenpox is usually a mild disease in healthy children”; “Chickenpox can cause serious complications”; “I think chickenpox is a disease serious enough to vaccinate against”; “I am worried about the side effects of chickenpox vaccination”; “Immunity from contracting chickenpox naturally is better than receiving the vaccine”; “It is better to have chickenpox when you are a child than when you are an adult”.

We then asked participants if they had personally seen complications of chickenpox and if this was in their own patients, their own child or other. If they indicated they had seen it in their own patients, we then asked them to estimate for every ten cases of chickenpox how many had complications. We also asked, “For every 10 parents you see for children’s vaccinations, how many would you say ask for chickenpox vaccination for their children?”.

Following this, we asked participants to indicate on a five-point Likert scale (from strongly agree to strongly disagree) their agreement with seven statements concerning the likely response from parents if/when the chickenpox vaccination is introduced in the UK and how they would feel about responding to parents, such as “Most parents will vaccinate their child against chickenpox” and “I would find it difficult to discuss the chickenpox vaccination with parents”.

All participants were then provided with some basic information about the infection and the vaccine (as we did for parents in Sherman et al., 2023). We also outlined the options for delivering the vaccination as follows: 1. Combining the chickenpox vaccine with MMR (MMRV); 2. Giving the chickenpox vaccine at the same time as MMR but as a separate injection (MMR+V); 3. Giving the chickenpox vaccine at an additional visit (V). Participants were asked which option they would prefer to deliver (they were given an ‘other’ option with free text as well). We asked a follow-up open-ended question asking them to explain the reasons for their choice. We then asked them to rank order the 3 options above in the order they thought parents would prefer them.

We then asked participants to indicate on a five-point Likert scale (from extremely likely to extremely unlikely) to what extent they thought each of 11 items (e.g., children already receiving too many injections; social media influence) might stop parents from having the chickenpox vaccine for their child/ren. Lastly, we asked to what extent they thought each of 9 items (e.g., reduction of chickenpox infection discomfort, advice from a well-informed healthcare professional) might encourage parents to accept the chickenpox vaccine. Both sets of items (adapted from Karafillakis et al., 2016) were supplemented with an ‘other’ option and an accompanying open-ended text box.

Two unrelated attention check questions were included as a quality control measure. Participants were excluded if they answered both of these questions incorrectly.

The study was reviewed and approved by Keele University’s Psychology Research Ethics Committee (reference: REC 0116).

## RESULTS

The survey was completed by a total of 91 participants aged between 26 and 65 years (mean=48.7, standard deviation=9.4) who were included in the data analysis. Participant characteristics are detailed in Table 1. Thirty-nine (42.9%) respondents reported having a child under the age of 18 and all reported that their child was up to date with their vaccines.

**Table 1.** Participant characteristics.

| Demographic questions | Level | *n* (%) |
| --- | --- | --- |
| Gender | Female | 88 (96.7) |
|  | Male | 3 (3.3) |
| Ethnicity | White | 83 (91.2) |
|  | Other ethnic groups | 7 (7.7) |
|  | Prefer not to say | 1 (1.1) |
| Religion | No religion | 43 (47.2) |
|  | Christian | 45 (49.5) |
|  | Other religion | 1 (1.1) |
|  | Prefer not to say | 2 (2.2) |
| Primary role | Lead nurse/nurse/nurse practitioner/trainee | 64 (70.3) |
|  | Health visitor | 10 (11.0) |
|  | Consultant/paediatric doctor | 6 (6.6) |
|  | Researcher | 3 (3.3) |
|  | GP/doctor | 2 (2.2) |
|  | Other | 6 (6.6) |
| Region | East Midlands | 18 (19.8) |
|  | East of England | 9 (9.9) |
|  | London | 8 (8.8) |
|  | North East & Cumbria | 1 (1.1) |
|  | North West | 6 (6.6) |
|  | Northern Ireland | 1 (1.1) |
|  | Scotland | 3 (3.3) |
|  | South East | 20 (22.0) |
|  | South West | 9 (9.9) |
|  | Wales | 2 (2.2) |
|  | West Midlands | 7 (7.7) |
|  | Yorkshire & the Humber | 5 (5.5) |
|  | Prefer not to say | 2 (2.2) |
| Have children under 18 | Yes | 39 (42.9) |
|  | No | 52 (57.1) |

### General vaccine attitudes

Participants attitudes towards vaccination in general are presented in Table 2.

**Table 2.**
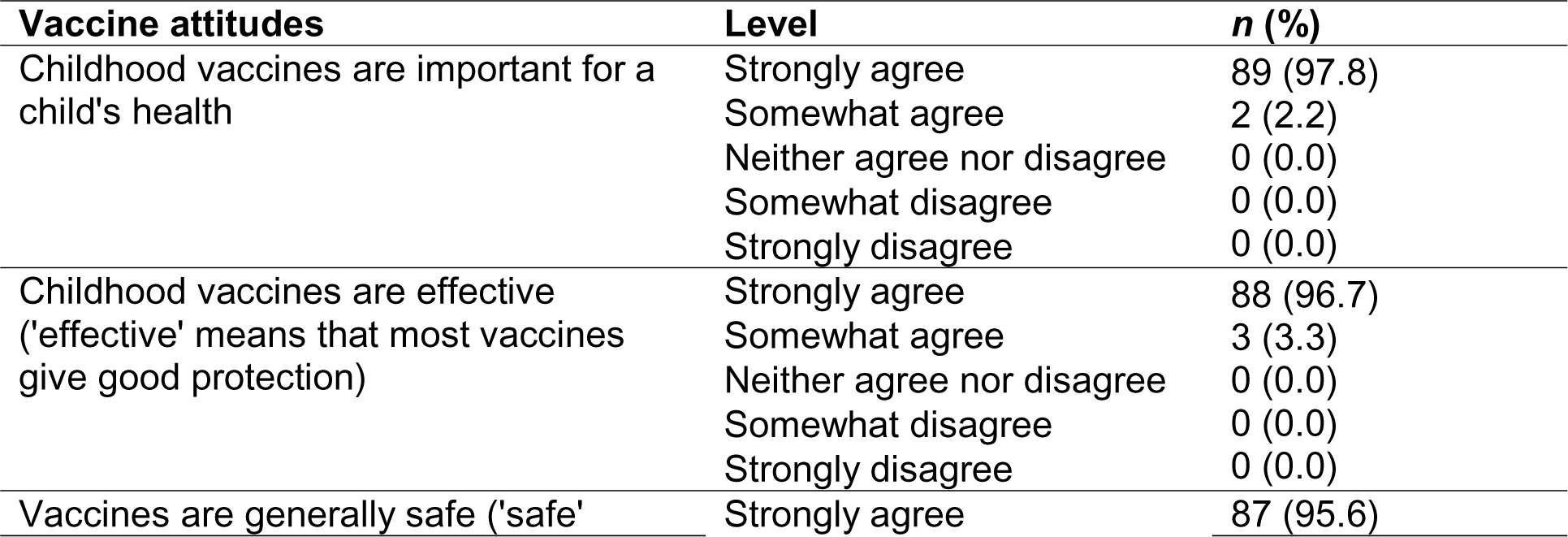
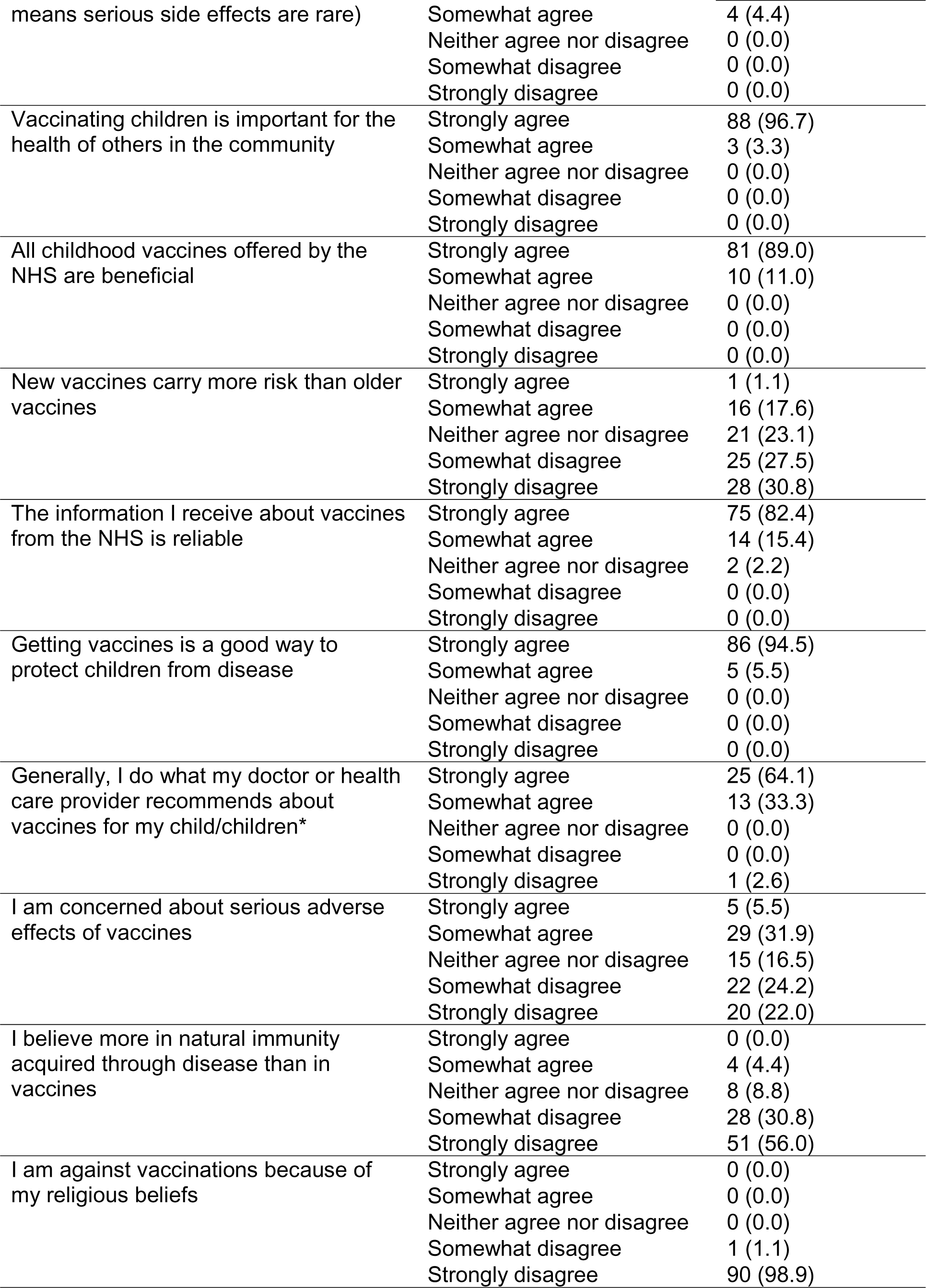
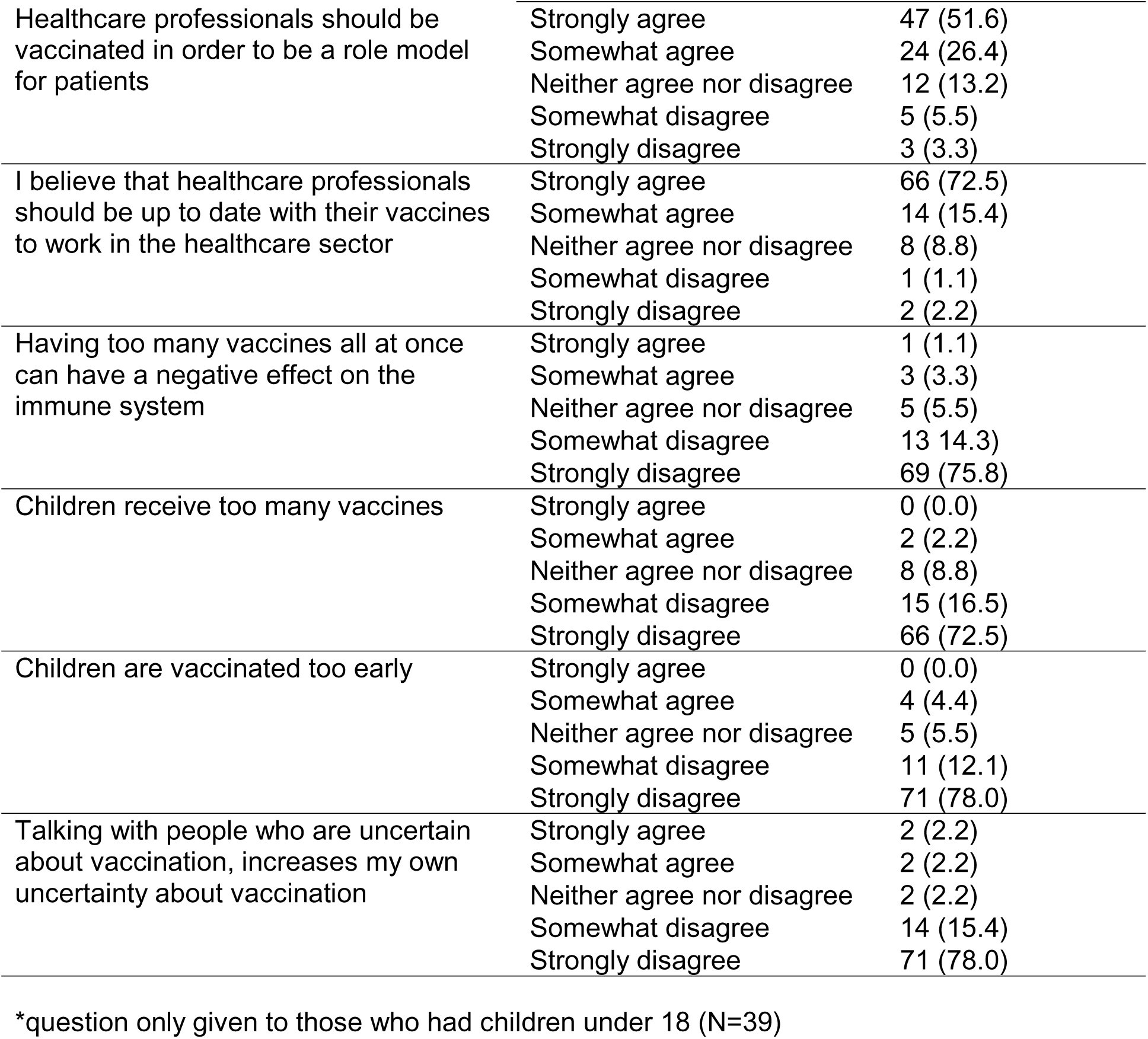
Healthcare professionals’ attitudes towards vaccination.

### Knowledge of Varicella disease

Participants’ knowledge about varicella are shown in Table 3.

**Table 3.** Healthcare professionals’ knowledge of varicella.

| Varicella knowledge | Level | n (%) |
| --- | --- | --- |
| Chickenpox generally has a mild disease course in healthy children | Strongly agree | 27 (29.7) |
|  | Somewhat agree | 50 (54.9) |
|  | Neither agree nor disagree | 6 (6.6) |
|  | Somewhat disagree | 7 (7.7) |
|  | Strongly disagree | 1 (1.1) |
| Chickenpox can cause serious complications | Strongly agree | 42 (46.2) |
|  | Somewhat agree | 30 (33.0) |
|  | Neither agree nor disagree | 6 (6.6) |
|  | Somewhat disagree | 8 (8.8) |
|  | Strongly disagree | 5 (5.5) |
| I think chickenpox is a disease serious enough to vaccinate against | Strongly agree | 26 (28.6) |
|  | Somewhat agree | 27 (29.7) |
|  | Neither agree nor disagree | 15 (16.5) |
|  | Somewhat disagree | 18 (19.8) |
|  | Strongly disagree | 5 (5.5) |
| I am worried about the side effects of chickenpox vaccination | Strongly agree | 1 (1.1) |
|  | Somewhat agree | 10 (11.0) |
|  | Neither agree nor disagree | 27 (29.7) |
|  | Somewhat disagree | 26 (28.6) |
|  | Strongly disagree | 27 (29.7) |
| Immunity from contracting chickenpox naturally is better than receiving the vaccine | Strongly agree | 8 (8.8) |
|  | Somewhat agree | 12 (13.2) |
|  | Neither agree nor disagree | 40 (44.0) |
|  | Somewhat disagree | 17 (18.7) |
|  | Strongly disagree | 14 (15.4) |
| It is better to have chickenpox when you are a child than when you are an adult | Strongly agree | 60 (65.9) |
|  | Somewhat agree | 24 (26.4) |
|  | Neither agree nor disagree | 6 (6.6) |
|  | Somewhat disagree | 0 (0.0) |
|  | Strongly disagree | 1 (1.1) |

### Personal experience of varicella

Forty respondents (44.0%) reported that they had personally seen complications from varicella. For those who indicated situations other than in their patients or their own child, these included among relatives or friends (n=5), personal experience (n=1) and seeing complications as a student working in adult intensive care (n=1). Respondents’ experiences of varicella are presented in Table 4.

**Table 4.** Responses to questions about experience of varicella.

| Question | Level | n (%) |
| --- | --- | --- |
| *Have you personally seen complications of chickenpox? | Yes, in my patients | 29 (31.9) |
|  | Yes, in my own child** | 6 (15.4) |
|  | Other | 7 (7.7) |
|  | No | 51 (56.0) |
|  | Prefer not to say | 0 (0.0) |
| For every 10 cases of chickenpox that you see, how many would you say have complications?*** | 1-2 out of 10 cases | 27 (67.5) |
|  | More than 2 out of 10 cases | 2 (5.0) |
| For every 10 parents you see for children's vaccinations, how many would you say ask for chickenpox vaccination for their children? | 1-2 | 40 (44.0) |
|  | 3-5 | 9 (9.9) |
|  | More than 5 | 2 (2.2) |
|  | None | 38 (41.8) |

\*Please select all that apply
\*\*denominator for this response is 39 (number of respondents with children under 18)
\*\*\*denominator for this response is 40 (number of respondents who have seen complications of chickenpox)

### Preferences for administration of varicella vaccine

Forty-three respondents (47.3%) indicated they would prefer to deliver the vaccine combined with MMR – the MMRV option, 25 (27.5%) preferred the MMR+V option and 20 (22.0%) preferred to give the vaccine at an additional visit (V). Three participants (3.3%) responded ‘other’, with one commenting that 4 vaccines are already a lot for a 12-month-old and the parent; another queried the relative risk of febrile convulsion for giving the vaccine at an additional visit, and the third indicated they did not want to give the varicella vaccine. Respondents were asked to provide an open-ended response explaining the rationale for their choice (see Table S4). The number of injections was the most frequently identified reason among those who selected MMRV and those who selected V. For those who selected MMR+V, the risk of febrile convulsions was the most frequently cited reason. Regarding likely parental preferences, 29 (39.2%) respondents indicated they thought parents would prefer the V option, 25 (33.8%) MMR+V and 20 (27.0%) MMRV. Predicted parental preferences for how the vaccine should be delivered are presented in Table 5.

**Table 5.** Predicted parental preference for administering vaccine (N=74)

| Item | Level | n (%) |
| --- | --- | --- |
| Combining the chickenpox vaccination with MMR (MMRV) | 1<sup>st</sup> choice | 20 (27.0) |
|  | 2<sup>nd</sup> choice | 22 (29.7) |
|  | 3<sup>rd</sup> choice | 32 (43.2) |
| Giving the chickenpox vaccination at the same time as MMR but as a separate injection (MMR+V) | 1<sup>st</sup> choice | 25 (33.8) |
|  | 2<sup>nd</sup> choice | 36 (48.6) |
|  | 3<sup>rd</sup> choice | 13 (17.6) |
| Giving the chickenpox vaccination at an additional visit (V) | 1<sup>st</sup> choice | 29 (39.2) |
|  | 2<sup>nd</sup> choice | 16 (21.6) |
|  | 3<sup>rd</sup> choice | 29 (39.2) |

### Likely response from parents

Respondents provided their level of agreement with statements about the likely response from parents if/when the chickenpox vaccination is introduced in the UK and their own perceived capability to address questions. The majority of respondents strongly or somewhat agreed that most parents will vaccinate their child against chickenpox (72.6%) and that they would feel confident to explain to parents the importance of vaccination against chickenpox (78.1%). The responses are presented in the Supplementary Materials.

### Potential barriers and facilitators to parents accepting varicella vaccine

Respondents were asked to identify the extent to which a range of issues might stop parents from having the chickenpox vaccine for their child/ren. The three potential barriers most frequently endorsed (strongly/somewhat agree responses) were perception that chickenpox is not serious, fear of vaccine side effects and social media influence. The responses are presented in the Supplementary Materials. Additionally, respondents were offered the opportunity to identify other barriers. Of 14 responses, five mentioned the influence of others such as friends, family, other parents, and playground gossip. Each of the following was also mentioned by one respondent: the anti-vaccination campaign, inconvenience of getting chickenpox, lack of clear government leadership and communication, lack of knowledge, older age for vaccination if the child has already had chickenpox infection, the idea of chickenpox parties so all children catch it when young and positive personal experience of this, concern about how long vaccine immunity will last and whether one is more likely to catch as an adult when immunity wanes, unable to get an appointment at their surgery due to staff shortages, and the influence of midwife/health visitor.

Respondents were asked to identify the extent to which a range of issues might encourage parents to accept the chickenpox vaccine. All of the potential facilitators were highly endorsed. The four most frequently endorsed were prior positive vaccination experience (96.7%), being aware that the vaccination will reduce the risk of rare but serious chickenpox complications (95.7%), advice from a well-informed healthcare professional (95.6%) and positive attitude towards vaccination from healthcare professional (95.6%). The responses are presented in the Supplementary Materials. Additionally, respondents were offered the opportunity to identify other facilitators. Of the 13 responses, five mentioned the influence of friends, families and other mothers, five the role of social and other media, with two highlighting positive messaging, two that it would mean parents would not have to take time off work with a sick child, one mentioned dispelling chickenpox parties and one knowing it has been introduced in other countries to good effect.

## DISCUSSION

To our knowledge this is the only contemporary survey of UK healthcare professionals’ views about introducing varicella vaccine to the routine childhood vaccination schedule. Although most participants recognised chickenpox to be a generally mild infection, they were also aware that it could result in serious complications. Only 58% of respondents strongly agreed or agreed that chickenpox is serious enough to vaccinate against.

We found that almost half of the HCPs in our survey would prefer to administer the varicella vaccine combined with MMR – the MMRV option, while the rest were almost evenly split between preferring the MMR+V option and giving the vaccine at an additional visit (V). A small number of HCPs responded that they did not favour an additional injection as the current schedule already includes four injections at 12 months. However, changes to the UK vaccination schedule due to be introduced (probably in 2025) when the current Hib/Men C booster will be removed (JCVI, 2022), will mean that even if a separate varicella vaccine was introduced, the total number of injections would still not exceed four. In contrast, more HCPs thought parents would prefer the V option, followed by MMR+V, with fewest respondents indicating they thought parents would prefer the MMRV option. This contrasts with our study of parents’ attitudes and preferences (Sherman et al, 2023) which suggested that parents would also prefer the MMRV option, with 65.6% of participants saying they would be extremely or somewhat likely to accept this option compared with 62.6% for V and 46.5% for MMR+V. In our survey, more than a fifth of our sample reported feeling either uncertain or disagreeing that they would feel confident explaining to parents the importance of vaccination against chickenpox. However, since the introduction of any vaccine in the UK is accompanied by information materials for parents and training for HCPs, this uncertainty will be addressed. Some of the responses in our survey highlighted areas that need to be addressed in these materials. For example, there was a spread of responses to the statement that new vaccines carry more risk than older vaccines, with nearly a fifth of respondents agreeing with this statement and more than a third of respondents indicated they were concerned about serious adverse effects of vaccines. Varicella vaccine has been used widely in many countries for decades, which should prove reassuring for those HCPs with concerns about the safety of apparently ‘new’ vaccines. Additionally, as there was some uncertainty about the severity of chickenpox and its association with serious complications, this will require clarification. Interestingly, most of our sample believed that HCPs should be vaccinated as a role model for patients, and up-to-date with vaccines to work in the healthcare sector.

There is additional evidence from this survey of an appetite among parents for varicella vaccine, with more than a half of respondents reporting that parents have asked them for the vaccine for their children. The majority of respondents also endorsed the view that most parents will vaccinate their child against chickenpox if/when the vaccine is introduced to the UK routine immunisation schedule. Two of the main potential barriers chimed with those identified by parents in Sherman et al. (2023), specifically the perception that chickenpox is not serious and fear of vaccine side effects, underlining the information that parents will require to inform their vaccine decisions.

This survey of HCPs’ attitudes and preferences for varicella vaccination complements the recent survey by Sherman et al. (2023) exploring the parent perspective. While the sample in this study is relatively small, it provides important insights into how HCPs would prefer to administer the vaccine as well as likely training needs should the vaccine be introduced to the routine schedule.

### Transparency declaration

The authors affirm that the manuscript is an honest, accurate, and transparent account of the study being reported; that no important aspects of the study have been omitted; and that any discrepancies from the study as originally planned have been explained.

### Data sharing statement

Data are available here: https://osf.io/vejmz/?view_only=2ace6027f9b4499c88059d24a9c3fc63 (public link to follow acceptance)

### Licence Statement

I, the Submitting Author has the right to grant and does grant on behalf of all authors of the Work (as defined in the author licence), an exclusive licence and/or a non-exclusive licence for contributions from authors who are: i) UK Crown employees; ii) where BMJ has agreed a CC-BY licence shall apply, and/or iii) in accordance with the terms applicable for US Federal Government officers or employees acting as part of their official duties; on a worldwide, perpetual, irrevocable, royalty-free basis to BMJ Publishing Group Ltd (“BMJ”) its licensees.

## Data Availability

The data and questionnaire are available online (https://osf.io/vejmz/?view_only=2ace6027f9b4499c88059d24a9c3fc63 [peer review link to be updated on acceptance with public link]).

## SUPPLEMENTARY MATERIALS

**Table S1.** Likely response from parents to introduction of varicella vaccination.

| Likely parental responses | Level | n (%) |
| --- | --- | --- |
| Most parents will vaccinate their child against chickenpox | Strongly agree | 21 (23.1) |
|  | Somewhat agree | 45 (49.5) |
|  | Neither agree nor disagree | 16 (17.6) |
|  | Somewhat disagree | 9 (9.9) |
|  | Strongly disagree | 0 (0) |
| Parents will worry about the chickenpox vaccination being 'new' | Strongly agree | 15 (16.5) |
|  | Somewhat agree | 53 (58.2) |
|  | Neither agree nor disagree | 9 (9.9) |
|  | Somewhat disagree | 12 (13.2) |
|  | Strongly disagree | 2 (2.2) |
| Parents are generally supportive of vaccinating their children | Strongly agree | 36 (39.6) |
|  | Somewhat agree | 53 (58.2) |
|  | Neither agree nor disagree | 2 (2.2) |
|  | Somewhat disagree | 0 (0) |
|  | Strongly disagree | 0 (0) |
| Parents will be worried about having another vaccine for their child | Strongly agree | 14 (15.4) |
|  | Somewhat agree | 50 (54.9) |
|  | Neither agree nor disagree | 16 (17.6) |
|  | Somewhat disagree | 8 (8.8) |
|  | Strongly disagree | 2 (2.2) |
| I would feel confident to explain to parents the importance of vaccination against chickenpox | Strongly agree | 40 (44.0) |
|  | Somewhat agree | 31 (34.1) |
|  | Neither agree nor disagree | 9 (9.9) |
|  | Somewhat disagree | 10 (11.0) |
|  | Strongly disagree | 1 (1.1) |
| I would expect many questions from parents about the chickenpox vaccination | Strongly agree | 35 (38.5) |
|  | Somewhat agree | 43 (47.3) |
|  | Neither agree nor disagree | 8 (8.8) |
|  | Somewhat disagree | 5 (5.5) |
|  | Strongly disagree | 0 (0) |
| I would find it difficult to discuss the chickenpox vaccination with parents | Strongly agree | 1 (1.1) |
|  | Somewhat agree | 6 (6.6) |
|  | Neither agree nor disagree | 22 (24.2) |
|  | Somewhat disagree | 26 (28.6) |
|  | Strongly disagree | 36 (39.6) |

**Table S2.**
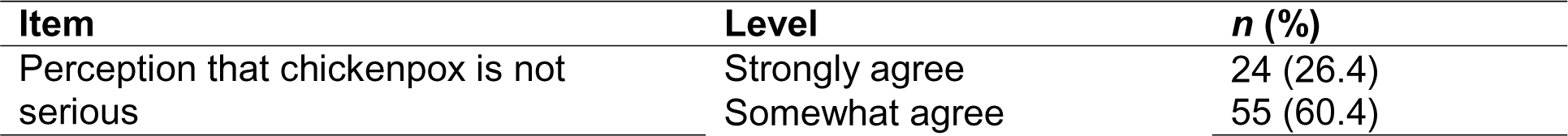
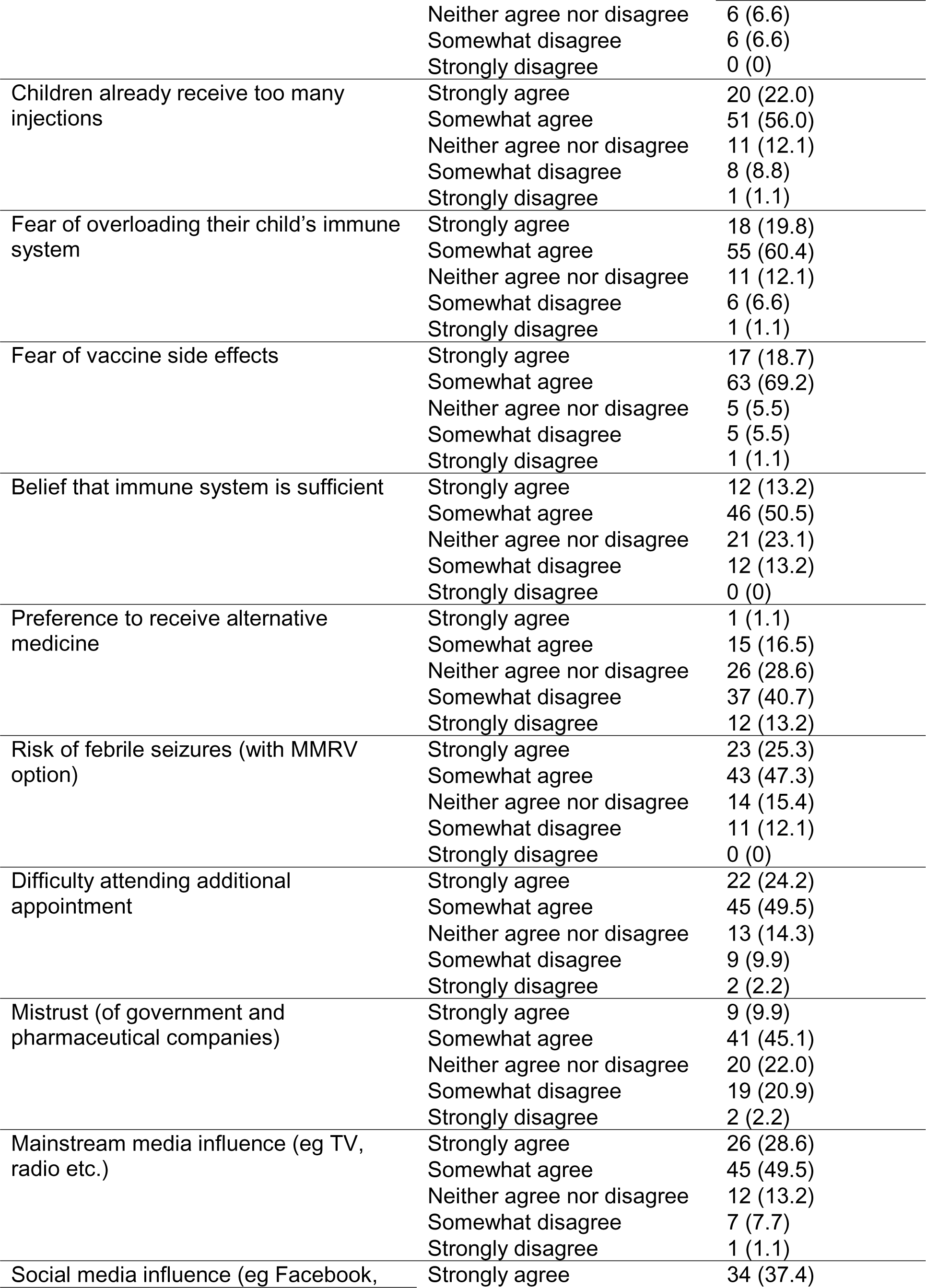
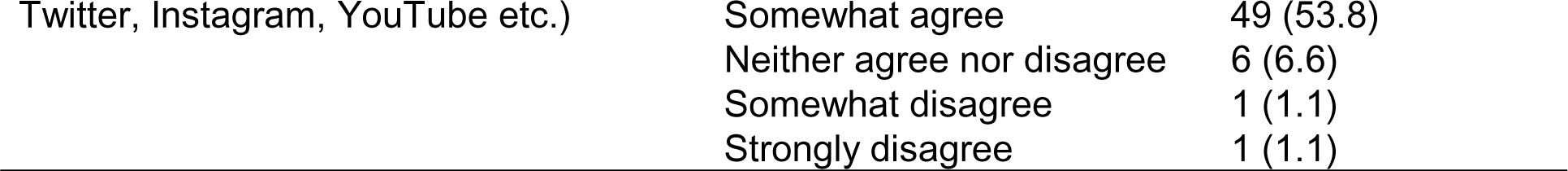
Potential barriers to parents accepting the varicella vaccination.

**Table S3.**
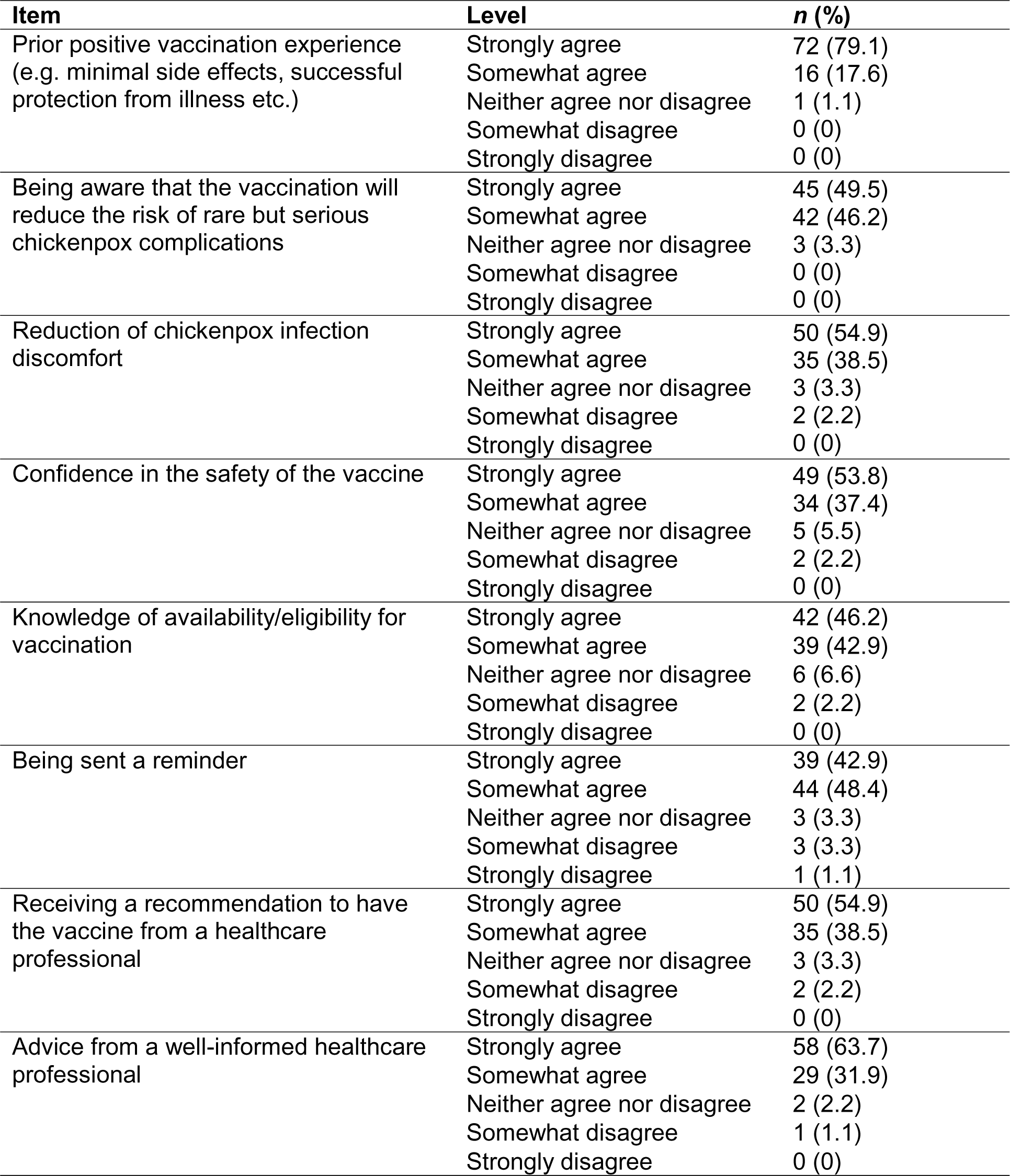
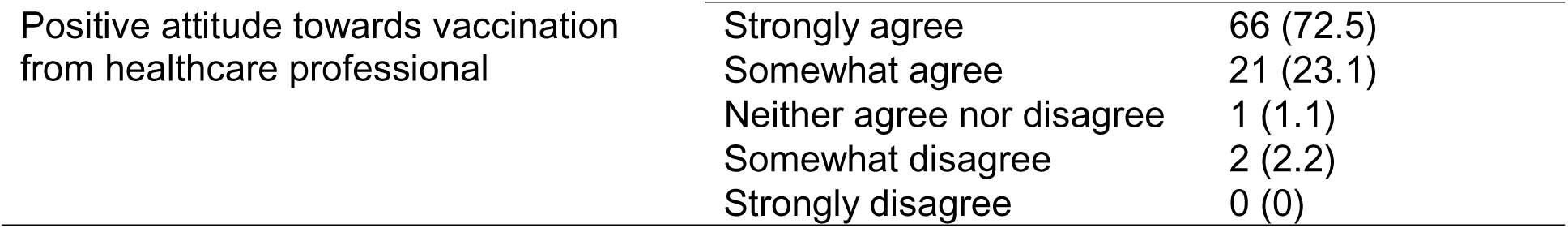
Potential facilitators to parents accepting the varicella vaccination.

| Item | Level | n (%) |
| --- | --- | --- |
| Prior positive vaccination experience<br>(e.g. minimal side effects, successful<br>protection from illness etc.) | Strongly agree | 72 (79.1) |
|  | Somewhat agree | 16 (17.6) |
|  | Neither agree nor disagree | 1 (1.1) |
|  | Somewhat disagree | 0 (0) |
|  | Strongly disagree | 0 (0) |
| Being aware that the vaccination will<br>reduce the risk of rare but serious<br>chickenpox complications | Strongly agree | 45 (49.5) |
|  | Somewhat agree | 42 (46.2) |
|  | Neither agree nor disagree | 3 (3.3) |
|  | Somewhat disagree | 0 (0) |
|  | Strongly disagree | 0 (0) |
| Reduction of chickenpox infection<br>discomfort | Strongly agree | 50 (54.9) |
|  | Somewhat agree | 35 (38.5) |
|  | Neither agree nor disagree | 3 (3.3) |
|  | Somewhat disagree | 2 (2.2) |
|  | Strongly disagree | 0 (0) |
| Confidence in the safety of the vaccine | Strongly agree | 49 (53.8) |
|  | Somewhat agree | 34 (37.4) |
|  | Neither agree nor disagree | 5 (5.5) |
|  | Somewhat disagree | 2 (2.2) |
|  | Strongly disagree | 0 (0) |
| Knowledge of availability/eligibility for<br>vaccination | Strongly agree | 42 (46.2) |
|  | Somewhat agree | 39 (42.9) |
|  | Neither agree nor disagree | 6 (6.6) |
|  | Somewhat disagree | 2 (2.2) |
|  | Strongly disagree | 0 (0) |
| Being sent a reminder | Strongly agree | 39 (42.9) |
|  | Somewhat agree | 44 (48.4) |
|  | Neither agree nor disagree | 3 (3.3) |
|  | Somewhat disagree | 3 (3.3) |
|  | Strongly disagree | 1 (1.1) |
| Receiving a recommendation to have<br>the vaccine from a healthcare<br>professional | Strongly agree | 50 (54.9) |
|  | Somewhat agree | 35 (38.5) |
|  | Neither agree nor disagree | 3 (3.3) |
|  | Somewhat disagree | 2 (2.2) |
|  | Strongly disagree | 0 (0) |
| Advice from a well-informed healthcare<br>professional | Strongly agree | 58 (63.7) |
|  | Somewhat agree | 29 (31.9) |
|  | Neither agree nor disagree | 2 (2.2) |
|  | Somewhat disagree | 1 (1.1) |
|  | Strongly disagree | 0 (0) |
| Positive attitude towards vaccination from healthcare professional | Strongly agree | 66 (72.5) |
|  | Somewhat agree | 21 (23.1) |
|  | Neither agree nor disagree | 1 (1.1) |
|  | Somewhat disagree | 2 (2.2) |
|  | Strongly disagree | 0 (0) |

**Table S4:** Reasons provided for choice of delivery preference.

| Item | MMRV | MMR+V | V |
| --- | --- | --- | --- |
| Number of injections | 26 | 3 | 9 |
| Reduce pressure on NHS (time, cost etc) | 12 |  | 1 |
| Increase uptake/reduce drop out | 11 | 3 |  |
| Number of visits | 11 | 2 |  |
| Reduce child distress | 8 |  |  |
| Reduce parental worry and hesitancy/parents prefer | 5 |  | 5 |
| Convenience | 3 | 1 |  |
| Men B precedent | 3 |  |  |
| Give parents a choice |  | 2 |  |
| Protection | 2 |  |  |
| Concern re impact on MMR | 1 | 7 | 2 |
| Separate at start of roll out | 1 | 1 |  |
| parent concerns re MMR | 1 |  | 6 |
| Easier to explain to parents/transparency | 1 | 2 | 1 |
| Less risk of side effects | 1 |  |  |
| Safe and effective | 1 |  |  |
| Already used abroad | 1 |  |  |
| Concern re parental reluctance | 1 |  |  |
| Concern re febrile seizures |  | 13 | 7 |
| Not enough info provided in this study |  | 1 |  |
| Identifiable side effects |  |  | 1 |
| Concern re fake news |  |  | 1 |
| Need time to counsel parents |  |  | 1 |
| Parents more anxious post COVID |  |  | 1 |

